# Performance and validation of a digital memory test across the Alzheimer’s Disease continuum

**DOI:** 10.1101/2024.06.18.24309112

**Authors:** Sofia Toniolo, Bahaaeddin Attaallah, Maria Raquel Maio, Younes Adam Tabi, Elitsa Slavkova, Verena Svenja Klar, Youssuf Saleh, Mohamad Imran Idris, Vicky Turner, Christoph Preul, Annie Srowig, Christopher Butler, Sian Thompson, Sanjay G. Manohar, Kathrin Finke, Masud Husain

**Author notes:** **Corresponding author:** Dr. Sofia Toniolo, Nuffield Department of Clinical Neurosciences, University of Oxford, New Radcliffe House, 1st Floor OX2 6GG Oxford.

## Abstract

Digital cognitive testing using online platforms has emerged as a potentially transformative tool in clinical neuroscience. In theory, it could provide a powerful means of screening for and tracking cognitive performance in people at risk of developing conditions such as Alzheimer’s Disease (AD). Here we investigate whether digital metrics derived from a tablet-based short-term memory task – “What was where?” Oxford Memory Task – were able to clinically stratify patients at different points within the AD continuum and to track disease progression over time. Performance of these metrics to traditional neuropsychological pen-and-paper screening tests of cognition was also analyzed. A total of 325 people participated in this study: 49 patients with subjective cognitive impairment (SCI), 57 with mild cognitive impairment (MCI), 63 with AD dementia and 156 elderly healthy controls (EHC). Most digital metrics were able to discriminate between healthy controls and patients with MCI and between MCI and AD patients. Some, including Absolute Localization Error, also differed significantly between patients with SCI and MCI. Identification accuracy was the best predictor of hippocampal atrophy, performing as well as standard screening neuropsychological tests. A linear support vector model combining digital metrics achieved high accuracy and performed at par with standard testing in discriminating between EHC and SCI (AUC 0.82) and between SCI and MCI (AUC 0.92). Memory imprecision was able to predict cognitive decline on standard cognitive tests over one year. Overall, these findings show how it might be possible to use a digital memory test in clinics and clinical trial contexts to stratify and track performance across the Alzheimer’s disease continuum.

## Introduction

Digital cognitive testing is being increasingly deployed as screening tool for patients at risk of developing Alzheimer’s Disease (AD), for recruitment in clinical trials and longitudinal follow-up performed remotely.^1,2^ Some tests can detect subtle signs of cognitive impairment that cannot be captured by standard clinical assessments.^3^ This makes their deployment potentially extremely valuable for large scale screening purposes in early phases of the disease when cognitive impairment is at subthreshold levels on current scales.^4^ Visual short-term memory (STM) tests have been extensively deployed in patients at risk of developing AD using digital platforms.^4,5^ Two types of paradigms have been used. The first involves change-detection tasks, in which participants indicate whether a change occurred or not in visual displays of differing numbers of items, in a binary correct/incorrect fashion.^6^ The second uses delayed-reproduction tasks, where participants are asked to reproduce features of the remembered item (e.g., its location, color or orientation) in a continuous response space. This allows modelling of the responses according to a resource model,^7^ where quantity (number of items held in memory) can be traded for quality (precision of recall of each item). The more items stored, the lower their precision in memory.

Mixture Modelling approach is a very influential computational model of how visual STM resources are allocated.^8^ In this model each response can be classified according to four different factors: probability of correctly identifying a target (**target detection**); erroneously placing an object at the location of another item in memory (**misbinding**); **random guessing** about the features of an item; **precision of memory** (probability distribution of the responses around the target).^8^ Previous analysis of performance on a delayed reproduction task, the “What was where?” Oxford Memory Task, demonstrated that people with a genetic risk factor for familial Alzheimer’s disease (FAD), such as carriers of the Presenilin-1 (PSEN1) mutation, exhibit higher misbinding rates, and that their recall correlated with the degree of hippocampal atrophy.^4^ Recent evidence using the same paradigm showed increased misbinding also in patients with sporadic, late-onset AD (LOAD).^9,10^ Whether this relates to hippocampal integrity is still unknown. Increased misbinding seems to be consistent across many different pathologies that target the hippocampus, including autoimmune limbic encephalitis,^11^ surgical resection due to epilepsy surgery,^12^ infectious encephalitis^13^ and anoxia.^14^ However, these are all relatively rare conditions, and extensive data on more common neurodegenerative conditions such as LOAD is missing.

Another important gap in the literature is the limited choice of digital outcomes used. Most previous studies have focused solely on misbinding rates, which is only one of a wide array of metrics that can be computed using digital delayed-reproduction tasks. This is important as there is evidence for selective disease-specific impairment in some cognitive metrics, whilst others are spared (e.g., increased misbinding but not guessing in patients with hippocampal pathology, and higher rates of guessing but not misbinding in patients with Parkinson’s Disease).^15^ Further, while we know that the degree of hippocampal atrophy relates to standard screening tests of cognitive function,^16^ and that delayed-reproduction visual STM metrics also show a good concordance with measures of hippocampal integrity,^4^ there are surprisingly little published data on head-to-head comparisons between digital STM metrics and commonly used clinical cognitive scales in predicting hippocampal integrity.

Longitudinal data on the temporal evolution of digital metrics, including misbinding, in patients at risk of developing AD, are also surprisingly scarce. At present, to our knowledge, only one study has reported on performance over time in a cohort of people with FAD, both symptomatic (n = 6) and presymptomatic (n = 23) gene mutation carriers.^5^ The authors used the “What was where?” Oxford Memory Task and found that identification accuracy declined over time only in symptomatic carriers. Moreover, localization error was greater the closer the presymptomatic individuals were to their estimated year of onset of dementia. Crucially, a standard delayed memory task, was only able to detect a significant difference between this group and controls one year later compared to localization error performance on the digital test. Despite these encouraging findings in FAD, performance of LOAD patients on this digital task has not been extensively characterized.

Here we report findings in a large group of individuals across the AD continuum, including people with subjective cognitive impairment (SCI), mild cognitive impairment (MCI) and established clinical AD dementia (AD). In this study, we sought to establish whether deficits on digital metrics can be detected before clinical diagnosis of AD dementia, and if these might also help to discriminate between clinical groups and elderly healthy controls (EHC). We tested individuals cross-sectionally and performed longitudinal assessment in a subset. Further, we investigated which among the digital metrics was the best predictor of cognitive decline longitudinally. The relationship of digital metrics to hippocampal integrity was also examined. Finally, a linear support vector machine^17^ was used to test the utility of digital metrics in classifying participants, and the resulting model was subsequently compared to one using a standard cognitive screening test.

## Material and methods

### Participants

325 participants were enrolled in the study: 49 people with SCI, 57 with MCI, 63 patients with AD dementia and 156 EHC. Patients were recruited from cognitive disorders clinics at the John Radcliffe Hospital in Oxford, United Kingdom and Friedrich-Schiller-Universität Klinik, Jena, Germany. SCI was defined according to the 2020 criteria from Jessen et al for subjective cognitive decline.^18^ MCI patients were classified according to Petersen’s criteria of 2014.^19^ Alzheimer’s disease dementia patients were defined as Alzheimer’s disease clinical syndrome according to the 2018 criteria by Jack et al^20^ and will be subsequently referred to as AD. From the Oxford cohort, 26/63 patients had plasma biomarkers analysis for AD biomarkers, with the results corroborating their diagnosis, based on clinical assessment, neuropsychological testing, magnetic resonance imaging (MRI) and fluorodeoxyglucose positron emission tomography (FDG-PET). All subjects underwent either brain computed tomography (CT) or MRI imaging and were excluded from the study if there was evidence of structural abnormalities not compatible with their clinical diagnosis. Elderly healthy subjects who reported any psychiatric or neurological illness or were on psychoactive drugs were excluded from the study. All participants had normal or corrected-to-normal vision acuity and no color blindness. A summary of participants’ demographics is presented in **Table 1**.

**Table 1.**
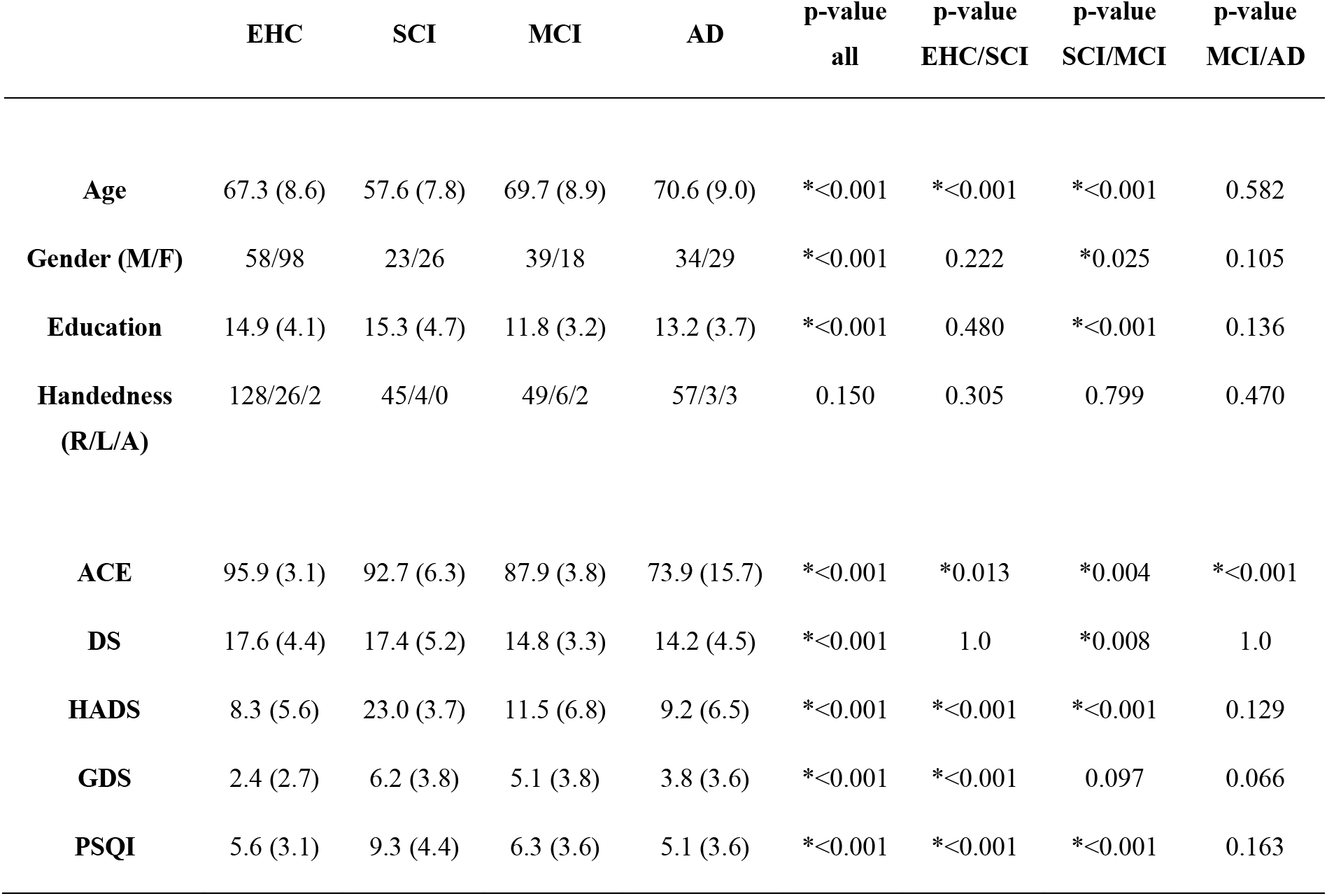
Demographics and tests. ACE=Addenbrookes Cognitive Examination-III; DS=Digit Span; HADS=Hospital Anxiety and Depression Scale; GDS=15-item Geriatric Depression Scale; and PSQI=Pittsburgh Sleep Quality Index. M = male, F = female. R = right-handed, L = left-handed, A = ambidextrous.

|  | EHC | SCI | MCI | AD | p-value<br>all | p-value<br>EHC/SCI | p-value<br>SCI/MCI | p-value<br>MCI/AD |
| --- | --- | --- | --- | --- | --- | --- | --- | --- |
| **Age** | 67.3 (8.6) | 57.6 (7.8) | 69.7 (8.9) | 70.6 (9.0) | *<0.001 | *<0.001 | *<0.001 | 0.582 |
| **Gender (M/F)** | 58/98 | 23/26 | 39/18 | 34/29 | *<0.001 | 0.222 | *0.025 | 0.105 |
| **Education** | 14.9 (4.1) | 15.3 (4.7) | 11.8 (3.2) | 13.2 (3.7) | *<0.001 | 0.480 | *<0.001 | 0.136 |
| **Handedness**<br>**(R/L/A)** | 128/26/2 | 45/4/0 | 49/6/2 | 57/3/3 | 0.150 | 0.305 | 0.799 | 0.470 |
| **ACE** | 95.9 (3.1) | 92.7 (6.3) | 87.9 (3.8) | 73.9 (15.7) | *<0.001 | *0.013 | *0.004 | *<0.001 |
| **DS** | 17.6 (4.4) | 17.4 (5.2) | 14.8 (3.3) | 14.2 (4.5) | *<0.001 | 1.0 | *0.008 | 1.0 |
| **HADS** | 8.3 (5.6) | 23.0 (3.7) | 11.5 (6.8) | 9.2 (6.5) | *<0.001 | *<0.001 | *<0.001 | 0.129 |
| **GDS** | 2.4 (2.7) | 6.2 (3.8) | 5.1 (3.8) | 3.8 (3.6) | *<0.001 | *<0.001 | 0.097 | 0.066 |
| **PSQI** | 5.6 (3.1) | 9.3 (4.4) | 6.3 (3.6) | 5.1 (3.6) | *<0.001 | *<0.001 | *<0.001 | 0.163 |

A subset of 60 people from the Oxford cohort took part in the longitudinal part of the study and completed the repeated assessment at 1 year. Of these 60 participants, there were 21 EHC, 15 SCI, 12 MCI, and 12 were patients with AD dementia. A summary of participants’ demographics is presented in **Supplementary Table S1.** The smaller numbers for the longitudinal dataset are due to the study being prematurely interrupted due to COVID-19 restrictions. While we have subsequently developed a fully remote, online version of this task,^21^ the data reported here are from a tablet version which required face-to-face administration.

138 participants from the Oxford cohort (EHC: n = 61, SCI, n = 31, MCI, n = 9, AD n = 37) agreed to a 3T structural MRI scan. Demographics and standard tests of cognition for this subsample are presented in **Supplementary Table S2**.

The study was performed in accordance with the ethical standards as laid down in the 1964 Declaration of Helsinki and its later amendments. Ethical approval was granted by the University of Oxford ethics committee (IRAS ID: 248379, Ethics Approval Reference: 18/SC/0448) and the local ethics committee in Jena. All participants gave written informed consent prior to the start of the study.

### Neuropsychological and behavioral test assessment

#### Cross-sectional assessment

The study protocol included a brief neuropsychological assessment and the “What was where?” Oxford Memory Task.^4,9,11^ The neuropsychological assessment included measures of global cognition, verbal short-term memory, depression, and sleep quality to rule out a potential neurodegenerative disorder or major depression in the elderly controls and for being able to compare performances at standard cognitive tests to our experimental paradigm. These included Addenbrooke’s cognitive examination (ACE-III, subsequently termed ACE),^22^ Digit Span (DS),^23^ Hospital Anxiety and Depression Scale (HADS),^24^ 15-item Geriatric Depression Scale (GDS)^25^ and Pittsburgh Sleep Quality Index (PSQI).^26^ Participants’ test scores are presented in **Table 1**.

A schematic of the “What was where?” Oxford Memory Task is shown in **Figure 1** (Panel A). Stimuli were presented on a black background and were chosen from a library of 196 fractals (http://sprott.physics.wisc.edu/fractals.htm), containing 49 different shapes of 4 colour variations each. Participants sat ∼30 cm in front of a tablet (either iPad or Android), yielding 2.3° of visual angle. Stimuli were calibrated using the dimension on the screen to ensure matching of stimuli properties across different tablet models. A fully remote online version of this task, available for computers, laptops, tablets and phones, has been subsequently developed (https://oxfordcognition.org/) and was used in subsequent studies.^21^

**Figure 1.**
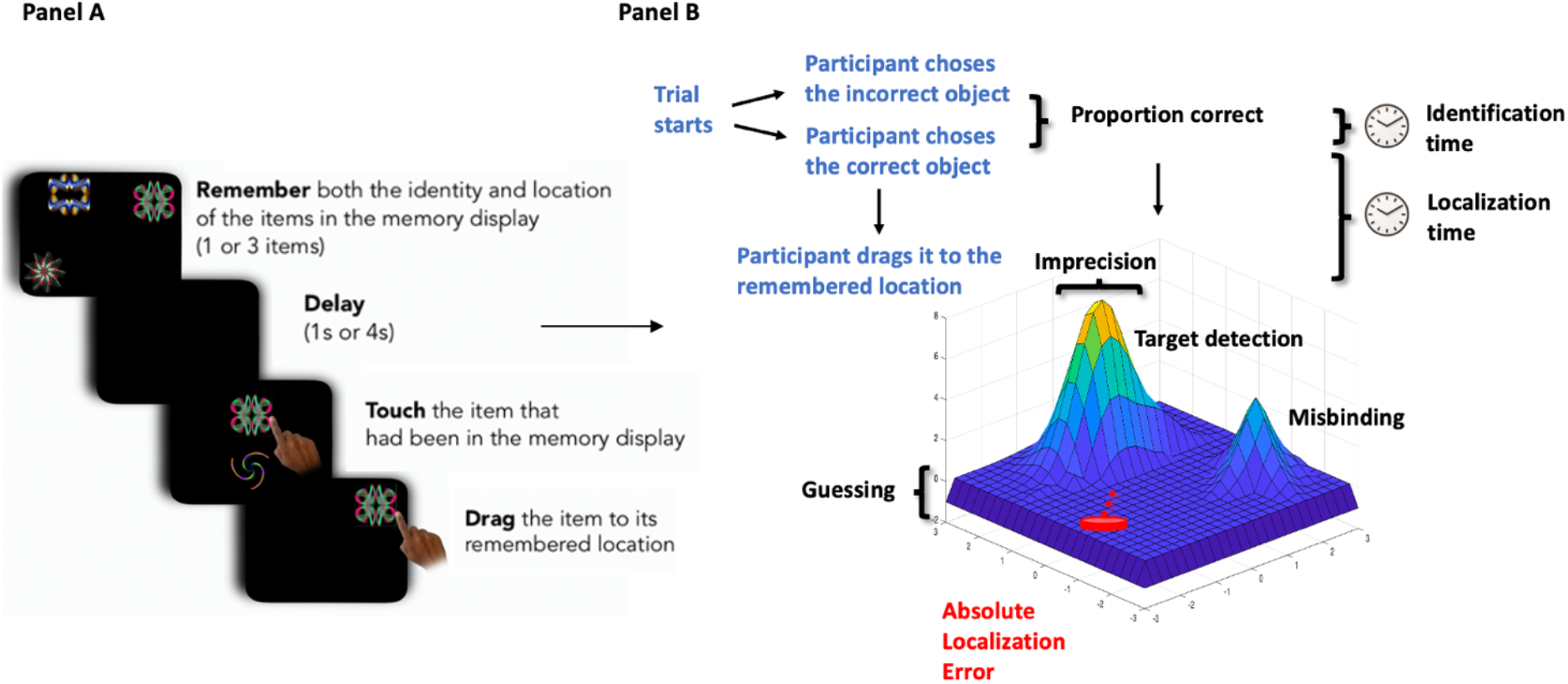
“What was where?” Oxford Memory Task (OMT) Panel A: This panel illustrates the task design. Participants were presented with either 1 or 3 fractals randomly distributed on the screen. After a 1 or 4 second delay two fractals appeared at the centre of the screen, one of which had appeared in the memory array whereas the other one was a distractor. Firstly, they needed to identify the object they had seen previously (‘what’), and then drag it back to its original location (‘where’). Panel B: The task provides four basic performance metrics: Identification accuracy; Absolute Localization Error; Identification time and Localization time. In addition, using the Mixture Model, it is possible to assess levels of Target detection, Misbinding, Guessing and Imprecision.

In different trials, participants were presented with either 1 or 3 fractals located randomly on the screen. They were asked to remember the identity of the fractals (‘what’), and their locations (‘where’). After a delay of either 1 or 4 seconds, two fractals appeared at the center of the screen along the vertical axis. One of these had appeared in the memory array (target) whereas the other one was a foil, which had not been shown in the current trial. Participants were required to touch the target and drag it to its original location. They were instructed to be as precise as possible, thus allowed to move the item and adjust its location as many times as they needed. This design yielded four different conditions (1 and 3 items, 1 and 4 seconds). Each participant performed a practice block of 8 trials followed by three test blocks of 40 trials, with a total of 120 test trials (10 trials x 4 conditions x 3 blocks). The order of the trials in a block was randomly chosen for each block. The locations of the fractals were determined by a MATLAB script (MathWorks, Inc) in a pseudorandom manner.

Overall, this task allowed to extract different working memory metrics (see **Figure 1, Panel B**), including (**basic metrics**):

- **Identification Accuracy**: the proportion of trials in which participants correctly identified the target.
- **Absolute Localization Error**: the Euclidian distance from the center of original item location to the center of participant’s response location.
- **Identification Time**: the time in seconds taken to identify the correct object.
- **Localization Time**: the time in seconds to drag the chosen object to its remembered location.

The Mixture Model of working memory by Bays et al^14^ was then fitted to the data to unravel the differential contribution of memory errors to our dataset. Model fitting was achieved using a permutation approach, where per each single trial, we calculated the distances between the response location and the location of:

1. Target.
2. Distractor (non-target in that trial).
3. A distractor taken from a randomly chosen trial.

Depending on which of these distances was the shortest, the response was either counted as target (1), distractor (2, i.e., misbinding) or random response (3, i.e., uniform guessing). We repeated this procedure 5000 times per trial, introducing a distractor from a randomly chosen trial each time. This procedure allowed us to calculate proportions for these three sources of response per trial (absolute amount of response type/5000). The introduction of a distractor that was randomly chosen from another trial allowed us to differentiate whether an error was systematically linked to the very specific trial’s distractor or whether it could be accounted for even by a randomly chosen distractor that was not present at trial. Importantly, these metrics were calculated uniquely on trials where an object was correctly identified. This approach has been previously published using this task and subsequent variations.^9,27^

The following metrics (**mixture model metrics**) were therefore calculated:

- **Target detection:** the probability of correctly identifying the target.
- **Misbinding**: the probability of mislocalizing a correctly identified item to the remembered location of another item in the memory array.
- **Guessing**: the probability of random guessing responses.
- **Imprecision**: the width of the distribution of the responses around the target.

#### Longitudinal assessment

At both visits all subjects completed the “What was where?” Oxford Memory Task, the ACE-III and DS. The summary of demographics and test scores can be found in **Supplementary Table S1.** Group differences in demographics and test scores were calculated using the same principles of the cross-sectional assessment.

### MRI acquisition and analysis

T1-weighted volumetric MR brain images were acquired on a 3T Siemens Magnetom Verio syngo scanner using a magnetisation prepared rapid gradient echo (MPRAGE) protocol acquired in sagittal orientation (TR = 2000 msec, TE = 1.94 msec, TI = 880 msec, Flip angle = 8 degrees, FOV read = 256 mm, Voxel size = 1.0 x 1.0 x 1.0 mm). All images were reviewed by a trained neurologist to exclude the presence of remarkable macroscopic brain abnormalities not compatible with the original diagnosis. Hippocampal volumes (HV) were estimated using FSL-FIRST.^28^ For each participant, left, and right HV were calculated, and bilateral HV was computed. We also calculated whole brain volumes for each subject. We subsequently computed the head size corrected values for whole brain volumes and HV, using the scaling factor derived from SIENAX.^29^ When referring to HV and whole brain volumes throughout the article, only head-size corrected volumes have been used. Images were carefully visually inspected after each processing step.

### Statistical analysis

All analyses were conducted using MATLAB 2019a, R (version 3.5.2) and JASP (JASP team, 2022). Data visualization was conducted using software Grammar of graphics plotting in MATLAB R2018b (Gramm library)^30^ and RStudio (Version 1.1.463). Statistical significance was set as p < 0.05, two-tailed.

#### Cross-sectional analysis

A one-way ANOVA was used to compare subjects’ continuous variables in demographics and test scores, with Holm post-hoc correction amongst the four groups, while χ^2^ test was used to compare categorical variables between groups. Additionally, a cumulative measure for each metric wad derived by calculating the mean across the 4 conditions (1 item 1 second, 1 item 4 seconds, 3 items 1 second, 3 items 4 seconds). An ANCOVA, with age, gender, and education as covariates, with subsequent Holm post-hoc correction, was used to test differences across the groups on each cumulative metric while controlling for factors that differed across groups and could impact performance.

We also examined the effects of Set size and Delay across all groups (transdiagnostically), calculating a 2 (Set size: 1 item, 3 items) x 2 (Delay: 1 second, 4 seconds) ANOVA for each of the digital working memory metrics. Effect size was quantified using Eta Squared (η2), defined as large (η2 > 0.14), medium (η2 > 0.06) or small (η2 > 0.01).^31^

#### Longitudinal analysis

A 4 (Group) x 2 (Session) ANCOVA, with age, gender and education as covariates, with subsequent Holm post-hoc correction, was used to test differences across the groups and sessions on each cumulative metric. Additionally, we tested whether the baseline values extracted from our tests could be used to predict cognitive decline over one year at standard neuropsychological tests. To this end, we calculated the change between ACE scores from the second visit and the first visit, and looked at whether any of the metrics was able to predict cognitive decline after 1 year. We ran linear regression for each metric separately and compared the results with the Cocor package in RStudio.^32^

#### MRI analysis

A generalized linear model was used to study correlations between neuropsychological measures and hippocampal volumes, while correcting for age, gender and education. Comparisons between regression coefficients were compared using the Cocor package in RStudio.^32^

#### Linear support vector machine

A linear support vector machine classifier (Classification learner, Matlab) was used to test the performance of a model including age, gender, education and the eight digital metrics derived from the “What was where?” Oxford memory Task (OMT), in discriminating between the four different diagnostic groups (EHC, SCI, MCI and AD), and a competing model using age, gender, education, and ACE scores, also derived from the same dataset.

## Results

### Demographics and standard neuropsychological tests

Age, gender, and education were not matched across groups, and therefore were included as covariates when comparing group performances (**Table 1**). As expected, ACE scores were statistically significantly different across groups, with AD showing the lowest scores, followed by MCI, then SCI and finally EHC. DS scores were not different between EHC and SCI but declined significantly in MCI and AD patients. Patients with SCI scored higher on questionnaires of depression and quality of sleep compared to the other groups.

### Cross-sectional analysis

All digital metrics were able to discriminate between the groups, with high effect size for a between-group difference (**Table 2**, column ALL, **Figure 2**). They were able to discriminate between EHC and AD and SCI and AD (**Table 2**, columns EHC/AD and SCI/AD). No metric was able to distinguish between EHC and SCI, which highlights the fact that despite these patients’ complaints, they perform within normal range on the visual STM test used here. On the other hand, compared to healthy controls, patients with MCI showed lower Identification Accuracy, had higher Absolute Localization Error rates, lower rates of Targets detected, higher amount of Guessing, higher rates of Misbinding, and higher memory Imprecision.

**Figure 2.**
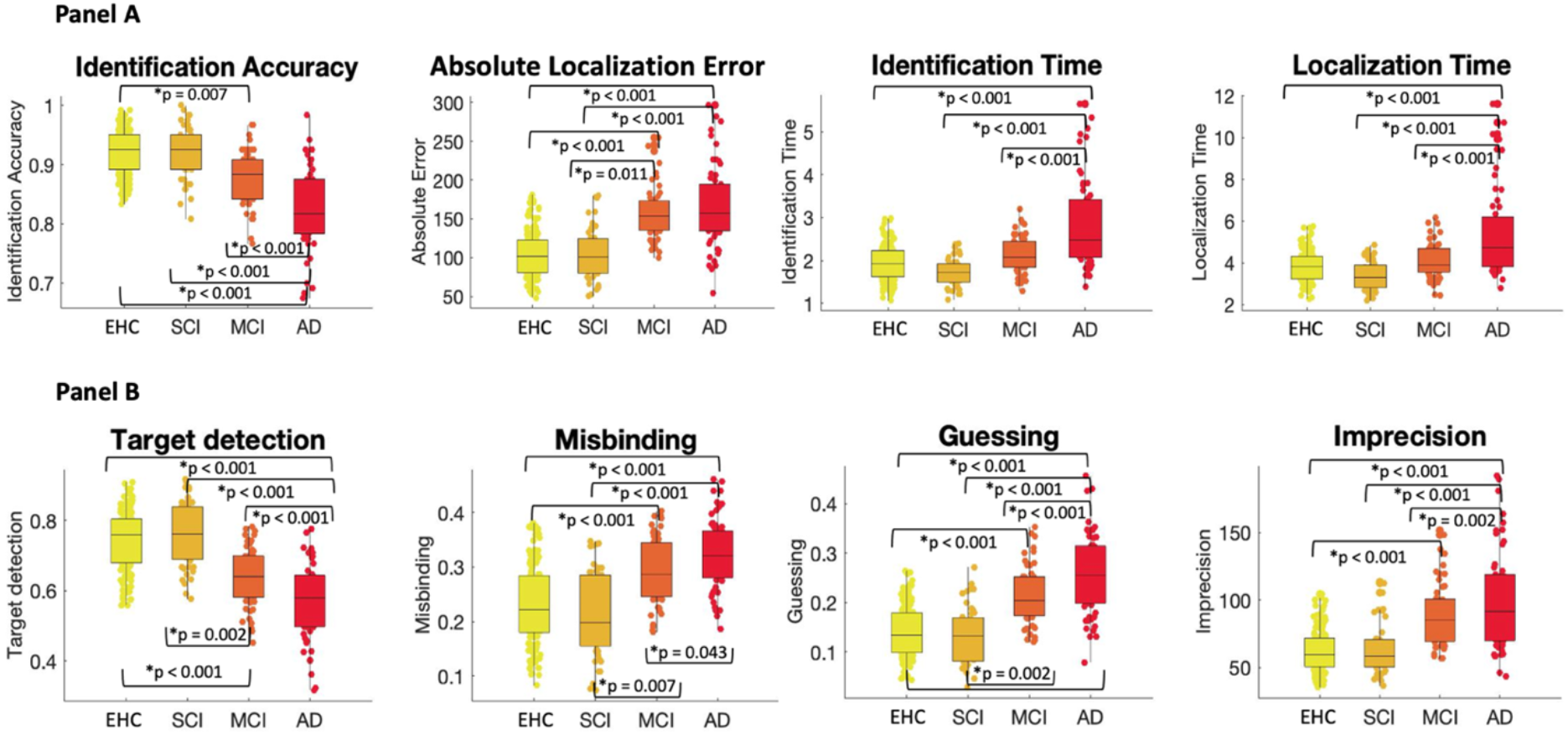
Cross-sectional analysis. Panel A: between-group analysis for basic performance metrics. Panel B: between-group analysis for mixture model metrics. An ANCOVA, with age, gender, and education as covariates, with subsequent Holm post-hoc correction, was used to test differences across the groups.

**Table 2.**
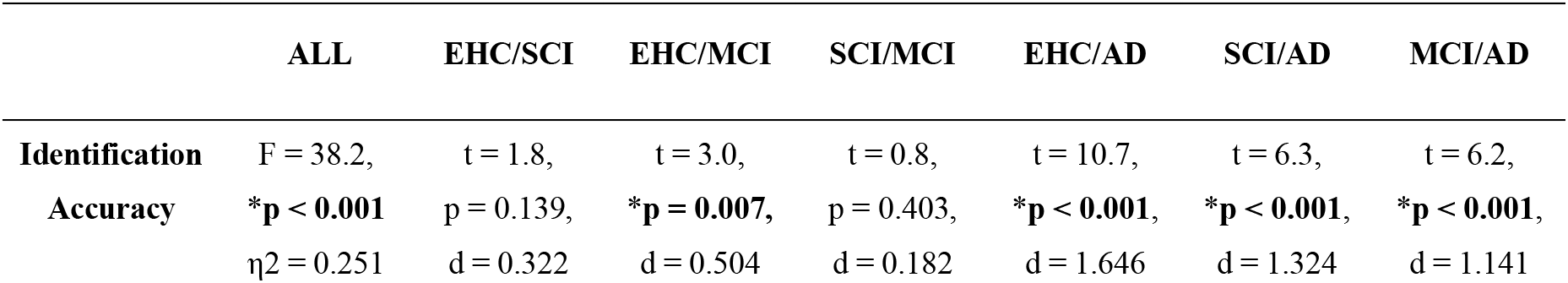
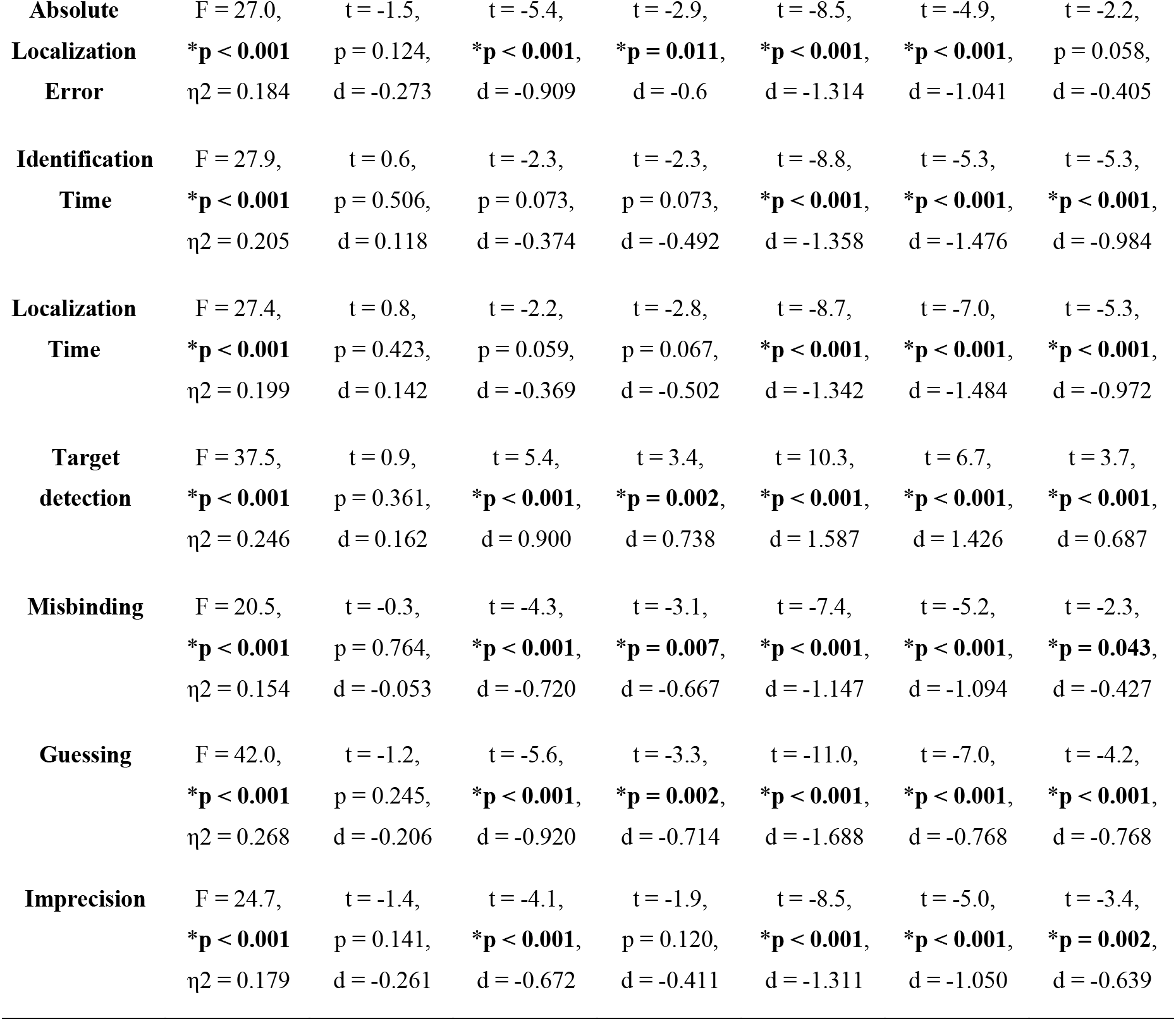
Summary results from the between-group analysis.

Absolute Localization Error, Target detection, Misbinding and Guessing were also able to discriminate between people with SCI and MCI (**Figure 2**, **Table 2**, column SCI/MCI). However, the precision of their memory recall (Imprecision), as well as the proportion of correctly identified items (Identification Accuracy) were not different between those groups. Absolute Localization Error was the only metric that was not able to track disease progression between patients with MCI and AD dementia, probably because it was already quite high in patients with MCI (**Figure 2** Panel A, **Table 2**, column MCI/AD). In the comparison between MCI and AD dementia, Misbinding was significantly higher in patients with AD dementia, but in this sample the metric had the lowest of all effect sizes (**Figure 2** Panel B, **Table 2**, column MCI/AD).

There was a main effect of Set size, of large magnitude (η2 > 0.14) in all metrics, except Imprecision, where it was of medium magnitude (η2 > 0.06), (**Supplementary Table S3 and S4**, **Figure S1 and S2**). Set size for Misbinding was not included because it cannot be computed for 1 item, as at least two objects are required for misbinding to occur. All metrics, except for Misbinding, also showed an effect of Delay, which however was of small (η2 > 0.01) to medium (η2 > 0.06) magnitude. Identification time, Identification accuracy, Absolute Localization Error and memory Imprecision also showed a significant Set Size by Delay interaction, which was however of small magnitude (η2 > 0.01), with a synergistic impact of higher number of items and longer delays determining worse memory performance.

### Longitudinal analysis

#### Group and Session effects

All metrics showed a significant effect of Group, with high effect size (η2 > 0.14), even if the sample size was much smaller compared to the bigger cross-sectional dataset (**Figure 3**, **Supplementary Table S5**). Only Localization Time showed a main effect of Session (F = 4.3, *p = 0.039, η2 = 0.024) (**Figure 3, Supplementary Table S5**). Absolute Localization Error and memory Imprecision showed a significant Group by Session interaction (**Figure 3, Supplementary Table S5**). Post-hoc analysis for Absolute Localization Error and Imprecision showed a significant difference between sessions only in the AD group (Absolute Localization Error: t = -3.120, *p = 0.031, d = - 1.274, Imprecision: t = - 4.064, *p = 0.002, d = -1.660).

**Figure 3.**
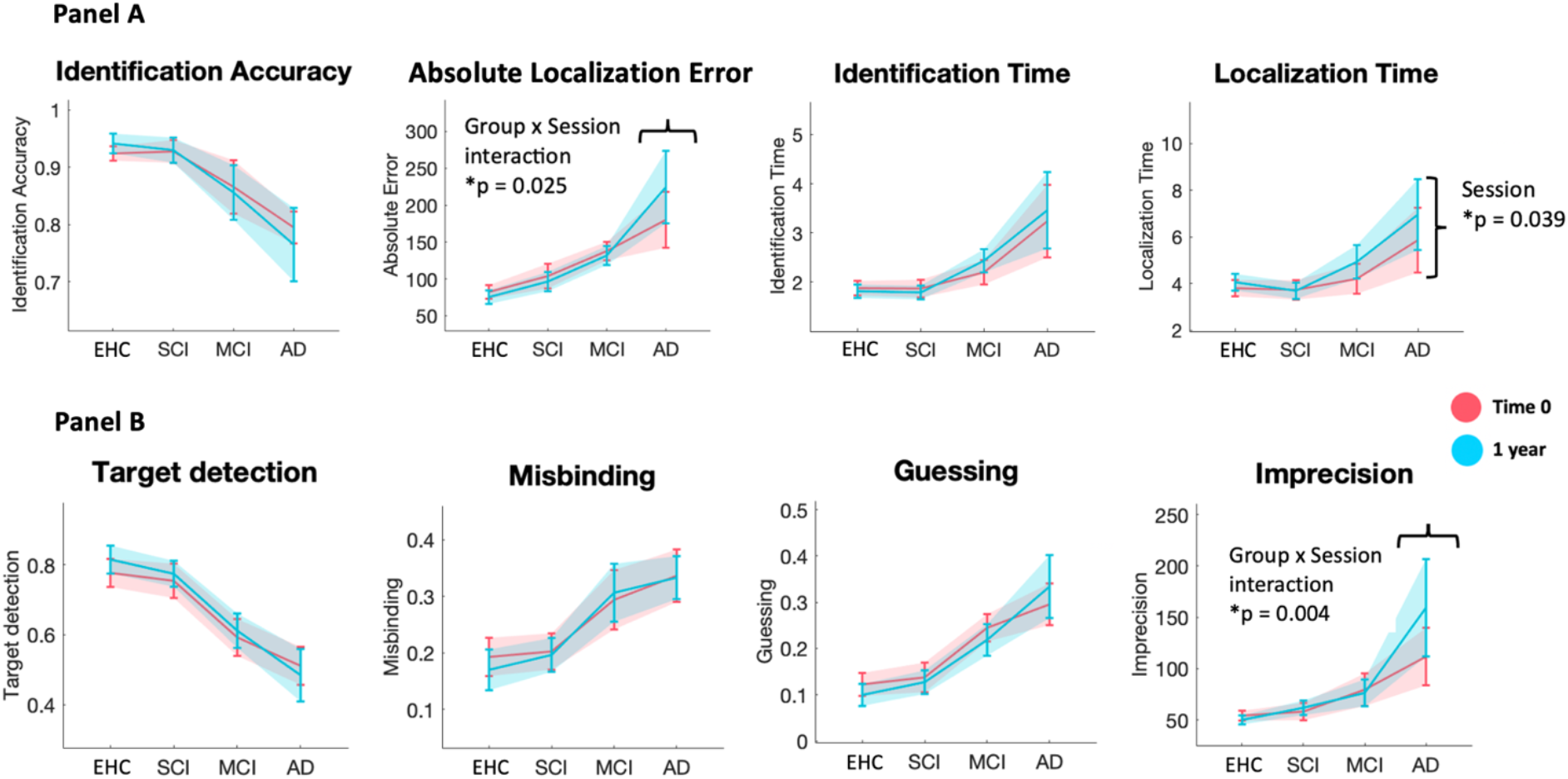
Longitudinal analysis. Panel A - longitudinal analysis of basic performance metrics. Panel B - longitudinal analysis of mixture model metrics. Baseline session (Time 0) in red, follow-up session after 1 year (1 year) in light blue.

As a comparison, we also computed the effects of Group and Session for the two neuropsychological tests used, the ACE and DS (**Supplementary Figure S3**). For ACE there was an effect of Group (F = 206.04, *p < 0.001, η2 = 0.812), an effect of Session (F = 10.16, *p = 0.002, η2 = 0.013), and a Group x Session interaction (F = 9.41, *p < 0.001, η2 = 0.037). For DS there was only an effect of Group (F = 6.67, *p < 0.001, η2 = 0.160), but no effect of Session or a Group x Session interaction. Post-hoc analysis was significant for ACE with respect to session for the AD group (F = 5.82, *p < 0.001), and for all groups comparisons (*p <0.001 except EHC vs SCI *p = 0.009), and for DS only for group comparisons between the AD group and the other groups (EHC vs AD *p <0.001, SCI vs AD *p = 0.003, MCI vs AD *p = 0.009).

#### Digital metrics in the prediction of cognitive decline

All metrics at baseline were able to independently predict cognitive decline after 1 year (**Supplementary Table S6**). The metric which performed the best in predicting a decline in ACE scores after 1 year was memory Imprecision, t = 5.5, *p < 0.001, R^2^ = 0.411. As a comparison, both ACE itself at baseline and hippocampal volume performed worse than memory Imprecision (ACE: h-test ACE, z = 6.34, *p < 0.001, Hippocampal volume: h-test, z = 5.74, *p < 0.001).

### Neuroimaging analysis

All metrics were able to independently predict hippocampal volume in the whole dataset (see **Figure 4**). As comparison, age had a smaller effect size compared to Identification accuracy in predicting hippocampal volume in the regression analysis (age: t = - 2.823, *p = 0.005, Identification accuracy, t = 3.251, *p = 0.001, h-test, z = - 4.3602, *p < 0.001). Moreover, Identification accuracy was the metric that was more tightly correlated with hippocampal volumes compared to the other metrics (h-test, z = 3.721, *p < 0.001), and performed as well as ACE (t = 3.761, *p < 0.001) in predicting hippocampal volume (h-test, z = - 0.190, p = 0.8494).

**Figure 4.**
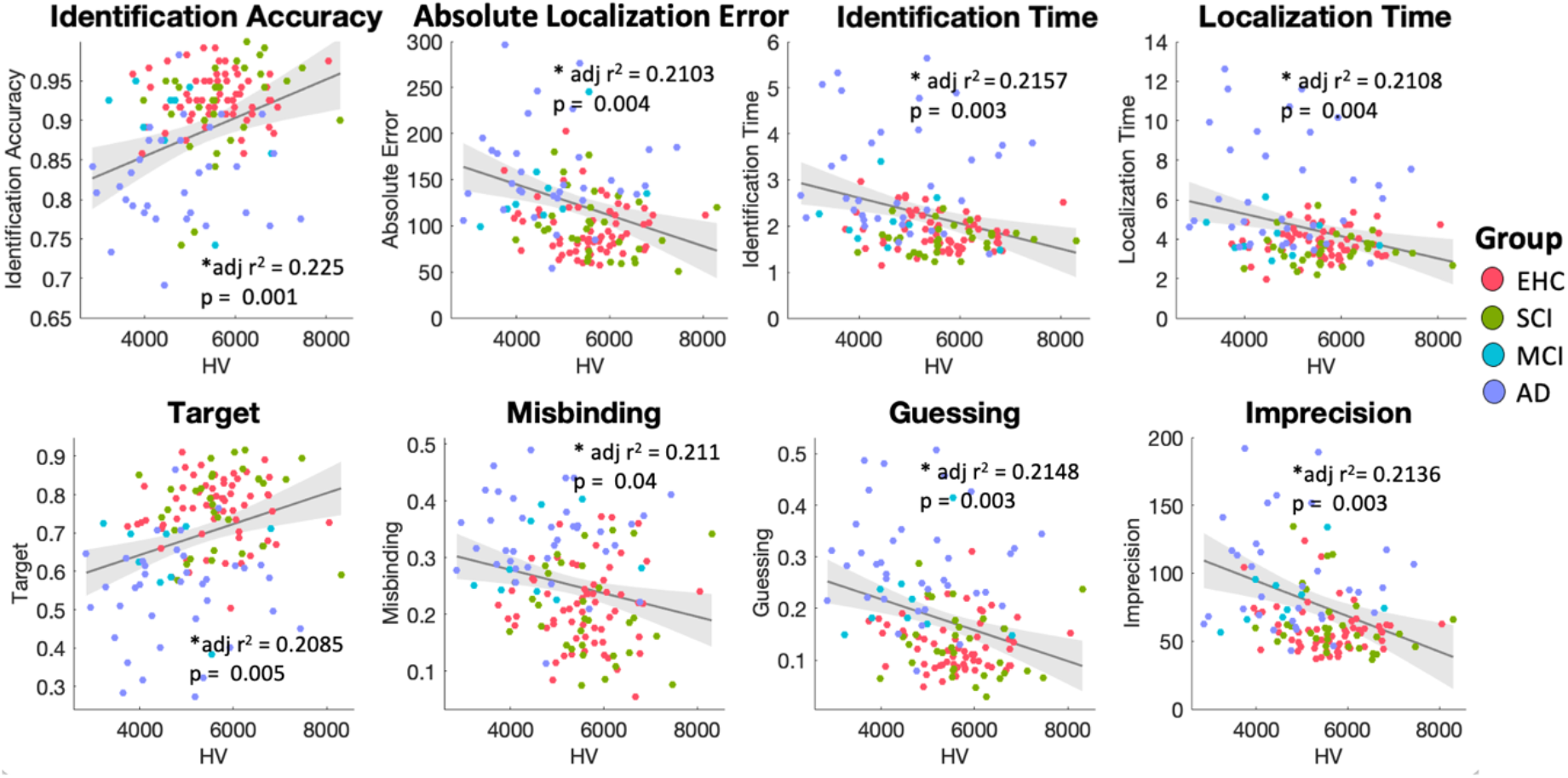
Correlations between hippocampal volumes and digital metrics. r^2^ represents the overall model fit, the p-value refers to the contribution of the metric to the regression. EHC in coral red, SCI in green, MCI in light blue, AD in violet. The regression line has been plotted for the whole dataset, in grey. HV = head-size corrected hippocampal volume, in mm^3^.

### Linear support vector machine

#### Overall classifier

For the model using the combination of the eight digital metrics (here labeled as OMT), overall accuracy was 61.8%, and the area under the curve (AUC) for predicting group classification was respectively 0.82 for healthy controls, 0.85 for patients with SCI, 0.80 for MCI, and 0.87 for AD dementia (**Figure 5**, Panel A – right, Panel B). In comparison, overall accuracy for the model including ACE was 71.4%, and the AUCs were respectively 0.92 for healthy controls, 0.83 for patients with SCI, 0.90 for MCI and 0.96 for AD dementia (**Figure 5**, panel A – left, Panel B).

**Figure 5.**
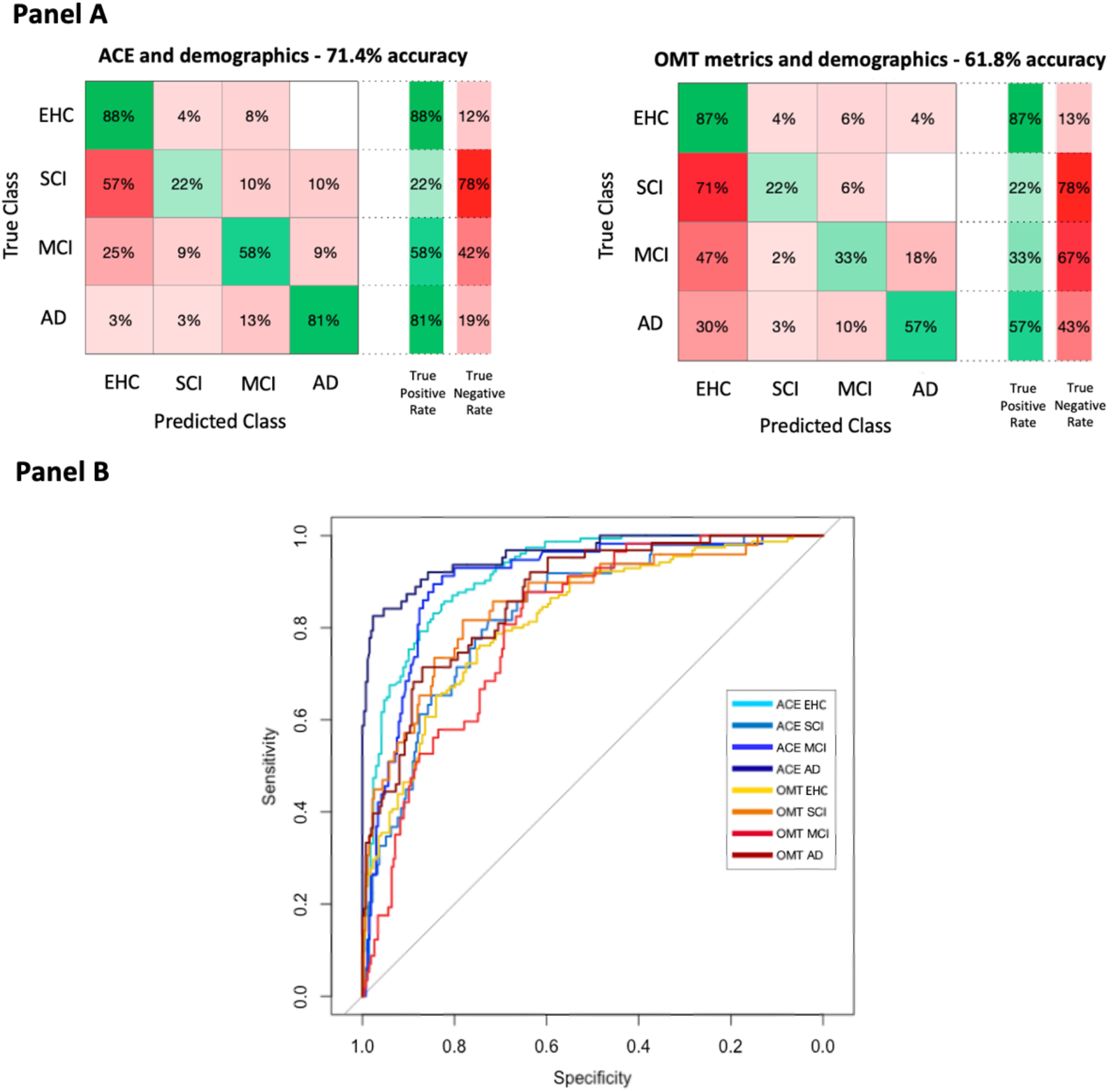
Linear support vector machine classifier – all groups. Panel A: Linear support vector machine classifier across all groups. The first model included ACE, age, gender and education, and the second all the eight OMT metrics, age, gender and education. Both models were tested for predicting group classification separately for EHC, SCI, MCI and AD. Panel B: Head-to-head comparison of ROC (receiver operating characteristic) curves for ACE and OMT models. Graded blue colours belong to ACE model: light blue = EHC, turquoise = SCI, blue = MCI, dark blue = AD. Graded red colour belong to the OMT model: yellow = EHC, orange = SCI, red = MCI, dark red = AD.

We computed pairwise comparisons using the DeLong method for comparing ROC curves^33^ between the model including OMT and the one including ACE for each group, which were respectively: EHC: Z = 4.83, *p < 0.001, SCI: Z = - 1.34, p = 0.179, MCI: Z = 3.81, *p < 0.001, AD: Z = 3.89, *p < 0.001. Therefore, the model using our digital metrics did not perform better than ACE in classifying healthy controls, patients with MCI and AD dementia, but was as good as the ACE in patients with SCI (**Figure 5**).

#### Pairwise classifier

Despite these multigroup classifiers being useful to blinded automatic diagnostic classifications, in a clinical setting we are less likely presented with a scenario when we have four different diagnoses that could fit the subject’s clinical profile. We therefore performed specific pairwise group comparisons where our metrics could potentially be clinically useful. As shown in **Figure 6**, the model including ACE and the one using OMT metrics perform similarly in the differential diagnosis between healthy elderly controls and patients with SCI (AUC ACE = 0.86, AUC OMT = 0.82, Z = 1.33, p = 0.182) and between SCI and MCI (AUC ACE = 0.91, AUC OMT = 0.92, Z = -0.757, p = 0.449). However, when examining patients with MCI and AD dementia, whilst the ACE performs extremely well, OMT metrics are not as good (AUC ACE = 0.91, AUC OMT = 0.75, Z = 2.84, *p = 0.004).

**Figure 6.**
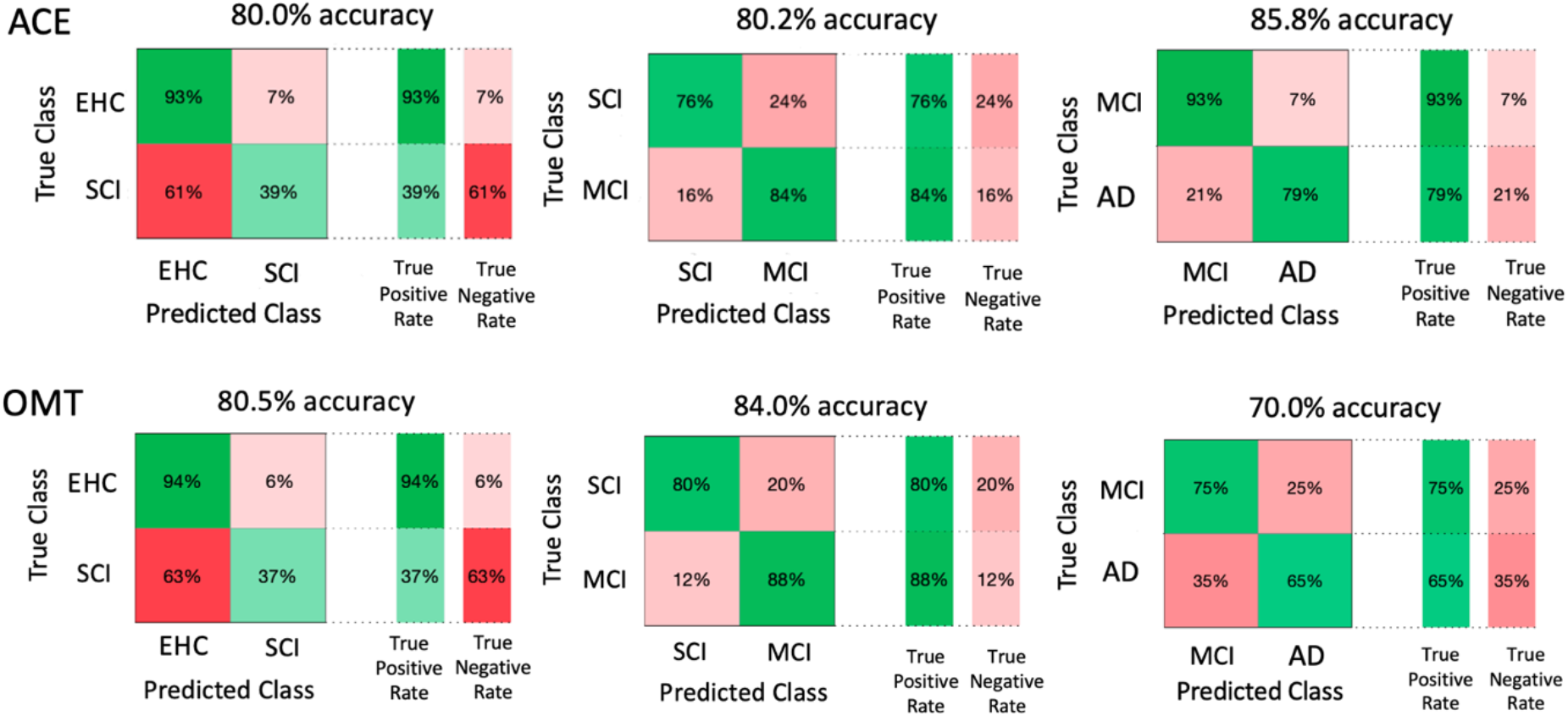
Linear support vector machine classifier – pairwise. The first model (ACE) includes ACE, age, gender and education, while the second (OMT) includes all eight OMT metrics, age, gender and education.

## Discussion

In this study we have shown that digital visual working memory metrics extracted by the “What was Where?” Oxford Memory Task are useful to detect early signs of cognitive impairment in a large cohort of subjects in the AD continuum. They are also able to detect disease progression cross-sectionally, between MCI and AD, but also longitudinally after 1 year. Identification accuracy seems the metric that best reflects the degree of underlying hippocampal atrophy, while Absolute Localization Error and memory Imprecision are useful to predict cognitive decline at standard tests of cognition over one year. Using a linear support vector machine classifier we found that these metrics seem to perform equally as good as standard tests of cognition such as the ACE in discriminating between healthy controls, patients with SCI and MCI, while they are not as accurate as ACE in later stages of the disease, such as in the comparison between MCI and AD dementia.

All digital metrics were able to discriminate between healthy controls and AD dementia patients, as well as between SCI and AD dementia (**Table 2**, **Figure 2**). However, no metric was able to distinguish between healthy controls and patients with SCI. This finding is consistent with data that show that the majority of people with a diagnosis of SCI will not subsequently develop AD dementia.^18^ Another important finding is that Absolute Localization Error, Target Detection, Misbinding and Guessing were all able to discriminate between patients with SCI and MCI. This might have potentially useful implications, as in clinical practice it can sometimes be difficult to distinguish between these groups on standard tests of cognition.

Both Identification accuracy and memory Imprecision did not differ significantly between healthy controls and patients with SCI, nor between SCI and MCI. However, there was a significant difference between healthy controls and the MCI group. In the comparison between MCI and AD, all digital metrics, except Absolute Localization Error, were significantly different. Absolute Localization Error seems to be sensitive to early stages of disease, as it was useful in discriminating between healthy controls and MCI, as well as between SCI and MCI. But beyond the MCI stage it did not deteriorate significantly further in patients with AD. Overall, Absolute Localization Error might be viewed as an ‘*early*’ digital marker because it seems to change more prominently in the early phases of cognitive impairment. Interestingly, it was the only marker which was able to highlight disease progression in asymptomatic mutation carriers in the FAD group studied by Pavisic et al,^5^ while in the same report Identification accuracy only declined in symptomatic patients. This further supports the idea that Absolute Localization Error might be considered as an early marker.

On the other hand, Identification time and Localization time might be viewed as ‘*late*’ markers, as in our sample they became significantly different only when patients reached the AD dementia stage. This is consistent with known effects of slowing of responses, e.g., slower reaction times (RT), in patients with AD dementia compared to age-matched elderly controls^34^ and patients with MCI.^35,36^ Evidence of slower reaction times in patients with MCI compared to age-matched controls has been more mixed, and whilst a recent metanalysis reported slower RT in this group,^37^ the results from our study do not support this finding. However, the investigations included in the metanalysis covered only two diagnostic groups, while here four groups were compared and the analyses were corrected for multiple comparisons, with a lower risk of bias but also possibly a reduced statistical significance in head-to-head differences.

Mixture model metrics including Target Detection, Guessing and Misbinding, were able not only to distinguish between a healthy status and MCI, but also to discriminate between patients with SCI and MCI and MCI and AD, so were useful metrics to provide an accurate stratification of patients and could be considered as both ‘*early*’ and ‘*late*’ markers.

Across groups, set size effects had a major impact on memory recall, showing an effect of large magnitude in all metrics, except for memory Imprecision, where it was of medium magnitude (**Supplementary Table S3 and S4**, **Figure S1 and S2**). This highlights how consuming STM capacity resources by adding items to remember results in lower performance, not only by reducing the precision with which the items are stored, but also causing slower responses, higher spatial localization error, lower percentages of correctly identified items and targets detected, and higher rates of random guessing and misbinding. Apart from Misbinding, all other metrics also showed an effect of delay, which however was of small to medium magnitude. This points towards the fact that remembering an increasing number of items has a much more detrimental impact on memory recall compared to time-dependent memory degradation. A significant effect of set size on memory performance is in line with what previously reported with the same paradigm in healthy controls and patients with sporadic AD and PD.^9,10^ However, the latter studies either did not have sufficient data to examine the effect of delay,^10^ or investigated only a limited subset of these metrics and included a large number of healthy controls but a very small number of patients with either AD or PD (n = 19 each).^9^ This study replicates these findings in a larger clinical cohort of patients with memory impairment.

Measuring changes in our cognitive metrics after one year revealed that they show different profiles: some tend to remain very stable over time, while others follow a similar trajectory as the decline in performance on the ACE (**Figure 3 and Supplementary Figure S3**). The metrics that best tracked individual changes in cognition over time were respectively Absolute Localization Error, but more so, memory Imprecision. Indeed, memory Imprecision performed very well in the longitudinal tracking of cognitive decline, so well that its baseline values outperformed baseline levels of ACE and hippocampal volumes (**Supplementary Table S6)**. One of the most interesting results was that despite a decline in cognition, indexed by a reduction in ACE scores, mixture model metrics (Target detection, Misbinding and Guessing) did not change over time. Why is that so? To answer this question, we need to bear in mind how these metrics are calculated. They do not reflect absolute values of errors, but they calculate the probability of a response belonging to correct target identification, misbinding and guessing, which all add up to 1, as they are relative to each other. Therefore, the most likely explanation is that the proportion of mistakes made, *relative to each other*, remains the same across time.

Previous evidence has pointed towards Misbinding as being the most important marker of hippocampal integrity across different clinical populations.^4,11^ While we could replicate that misbinding rates are indeed associated with hippocampal atrophy, this was not the best predictor among the digital metrics used for this experiment. Here, Identification accuracy stood out as the metric that most strongly correlated with hippocampal volume, performing equally as well validated standard neuropsychological test such as the ACE (**Figure 4**). It is also worth noting that these results were corrected for common confounding factors that could impact performance as well as hippocampal integrity, such as age, gender and levels of education. More importantly, Identification accuracy explained more variance compared to ageing in predicting hippocampal volume in this dataset, which is also encouraging.

Regarding head-to-head comparisons between digital metrics and standard tests of cognition, while ACE was overall superior to OMT metrics and better in the discrimination between MCI and AD dementia, digital metrics seem to perform similarly to traditional pen-and-paper tests when comparing healthy controls to SCI patients, or SCI to MCI, without requiring dedicated, time-consuming face-to-face appointments (**Figure 5 and 6**). This has potential useful clinical implications as this test can now be administered fully online and remotely.

There are some limitations to this study. Only 26/63 patients of the Oxford cohort had biological confirmation of AD biomarkers, which confirmed presence of amyloid and tau positivity. Whilst this sample is representative of patients seen in a memory clinic, subsequent studies should aim to validate these findings in a fully biomarker-validated cohort. Moreover, the longitudinal dataset had a limited number of participants, due to the study being interrupted in response to COVID-19 restrictions. Additionally, participants enrolled in the longitudinal dataset were all non-converters, i.e., none of them had their diagnosis changed at their follow-up visit after 1 year. A much larger longitudinal dataset is needed to replicate these findings and to study which metric might be a better marker of clinical conversion from early stages (e.g., SCI and MCI) to AD dementia.

## Data Availability

De-identified data supporting this study may be shared based on reasonable written requests to the corresponding author. Access to de-identified data will require a Data Access Agreement and IRB clearance, which will be considered by the institutions who provided the data for this research.

## Author contributions

**Toniolo Sofia**: conceptualization, participants’ recruitment, project administration, data collection and curation, behavioural and imaging analysis, data visualisation, writing.

**Bahaaeddin Attaallah**: data collection and curation, imaging analysis.

**Maria Raquel Maio**: project administration, data collection and curation, code development.

**Younes Adam Tabi**: code development, data collection and curation.

**Elitsa Slavkova**: project administration, data collection and curation.

**Verena Svenja Klar**: data collection and curation.

**Youssuf Saleh**: data collection and curation.

**Imran Idris**: data collection and curation.

**Vicky Turner**: data collection and curation.

**Christoph Preul**: data collection and curation.

**Annie Srowig**: data collection and curation.

**Steffen Joedecke:** data collection and curation.

**Christopher Butler:** participants’ recruitment.

**Sian Thompson:** participants’ recruitment.

**Sanjay Manohar:** conceptualization, participants’ recruitment, code development, software development, supervision.

**Kathrin Finke:** supervision, resources management, funding acquisition, participants’ recruitment.

**Masud Husain:** conceptualization, supervision, resources management, funding acquisition, participants’ recruitment, writing.

All authors read and approved the final manuscript.

## References

1. van der Flier WM, de Vugt ME, Smets EMA, Blom M, Teunissen CE. Towards a future where Alzheimer’s disease pathology is stopped before the onset of dementia. Nat Aging 2023 35. 2023;3(5):494–505. doi:10.1038/s43587-023-00404-2

2. Tsoy E, La Joie R, VandeVrede L, et al. Scalable plasma and digital cognitive markers for diagnosis and prognosis of Alzheimer’s disease and related dementias. Alzheimers Dement. 2024. doi:10.1002/ALZ.13686

3. Shibata K, Attaallah B, Tai XY, et al. Remote digital cognitive assessment reveals cognitive deficits related to hippocampal atrophy in autoimmune limbic encephalitis: a cross-sectional validation study. eClinicalMedicine. 2024;0(0). doi:10.1016/j.eclinm.2024.102437

4. Liang Y, Pertzov Y, Nicholas JM, et al. Visual short-term memory binding deficit in familial Alzheimer’s disease. Cortex. 2016. doi:10.1016/j.cortex.2016.01.015

5. Pavisic IM, Nicholas JM, Pertzov Y, et al. Visual short-term memory impairments in presymptomatic familial Alzheimer’s disease: A longitudinal observational study. Neuropsychologia. 2021;162. doi:10.1016/J.NEUROPSYCHOLOGIA.2021.108028

6. Parra MA, Abrahams S, Logie RH, Méndez LG, Lopera F, Della Sala S. Visual short-term memory binding deficits in familial Alzheimer’s disease. Brain. 2010. doi:10.1093/brain/awq148

7. Ma WJ, Husain M, Bays PM. Changing concepts of working memory. Nat Neurosci. 2014. doi:10.1038/nn.3655

8. Bays PM, Catalao RFG, Husain M. The precision of visual working memory is set by allocation of a shared resource. J Vis. 2009. doi:10.1167/9.10.7

9. Tabi YA, Maio MR, Attaallah B, et al. Vividness of visual imagery questionnaire scores and their relationship to visual short-term memory performance. Cortex. 2022;146:186–199. doi:10.1016/J.CORTEX.2021.10.011

10. Zokaei N, Sillence A, Kienast A, et al. Different patterns of short-term memory deficit in Alzheimer’s disease, Parkinson’s disease and subjective cognitive impairment. Cortex. 2020;132:41. doi:10.1016/J.CORTEX.2020.06.016

11. Pertzov Y, Miller TD, Gorgoraptis N, et al. Binding deficits in memory following medial temporal lobe damage in patients with voltage-gated potassium channel complex antibody-associated limbic encephalitis. Brain. 2013. doi:10.1093/brain/awt129

12. Finke C, Braun M, Ostendorf F, et al. The human hippocampal formation mediates short-term memory of colour-location associations. Neuropsychologia. 2008;46(2):614–623. doi:10.1016/J.NEUROPSYCHOLOGIA.2007.10.004

13. Olson IR, Moore KS, Stark M, Chatterjee A. Visual Working Memory Is Impaired when the Medial Temporal Lobe Is Damaged. J Cogn Neurosci. 2006;18(7):1087–1097. doi:10.1162/JOCN.2006.18.7.1087

14. Hannula DE, Tranel D, Cohen NJ. The Long and the Short of It: Relational Memory Impairments in Amnesia, Even at Short Lags. J Neurosci. 2006;26(32):8352. doi:10.1523/JNEUROSCI.5222-05.2006

15. Zokaei N HM. Working Memory in Alzheimer’s Disease and Parkinson’s Disease. Curr Top Behav Neurosci. 2019. doi:10.1007/7854_2019_103

16. Schuff N, Woerner N, Boreta L, et al. MRI of hippocampal volume loss in early Alzheimer’s disease in relation to ApoE genotype and biomarkers. Brain. 2009;132(Pt 4):1067–1077. doi:10.1093/BRAIN/AWP007

17. Cortes C, Vapnik V, Saitta L. Support-vector networks. Mach Learn 1995 203. 1995;20(3):273–297. doi:10.1007/BF00994018

18. Jessen F, Amariglio RE, Buckley RF, et al. The characterisation of subjective cognitive decline. Lancet Neurol. 2020;19(3):271. doi:10.1016/S1474-4422(19)30368-0

19. Petersen RC, Caracciolo B, Brayne C, Gauthier S, Jelic V, Fratiglioni L. Mild cognitive impairment: A concept in evolution. J Intern Med. 2014. doi:10.1111/joim.12190

20. Jack CR, Bennett DA, Blennow K, et al. NIA-AA Research Framework: Toward a biological definition of Alzheimer’s disease. Alzheimers Dement. 2018;14(4):535–562. doi:10.1016/J.JALZ.2018.02.018

21. Toniolo S, Zhao S, Scholcz A, et al. Relationship of plasma biomarkers to digital cognitive tests in Alzheimer’s disease. *Alzheimer’s Dement (Amsterdam*, Netherlands). 2024;16(2). doi:10.1002/DAD2.12590

22. Hsieh S, Schubert S, Hoon C, Mioshi E, Hodges JR. Validation of the Addenbrooke’s Cognitive Examination III in Frontotemporal Dementia and Alzheimer’s Disease. Dement Geriatr Cogn Disord. 2013;36(3-4):242–250. doi:10.1159/000351671

23. Wambach D, Lamar M, Swenson R, Penney DL, Kaplan E, Libon DJ. Digit Span. In: Encyclopedia of Clinical Neuropsychology. ; 2011. doi:10.1007/978-0-387-79948-3_1288

24. Skapinakis P. Hospital Anxiety and Depression Scale (HADS). In: Encyclopedia of Quality of Life and Well-Being Research. ; 2014. doi:10.1007/978-94-007-0753-5_1315

25. Yesavage JA, Brink TL, Rose TL, et al. Development and validation of a geriatric depression screening scale: a preliminary report. J Psychiatr Res. 1982;17(1):37–49. doi:10.1016/0022-3956(82)90033-4

26. Buysse DJ, Reynolds CF, Monk TH, Berman SR, Kupfer DJ. The Pittsburgh sleep quality index: A new instrument for psychiatric practice and research. Psychiatry Res. 1989. doi:10.1016/0165-1781(89)90047-4

27. Tabi YA, Maio MR, Fallon SJ, et al. Impact of processing demands at encoding, maintenance and retrieval in visual working memory. Cognition. 2021;214. doi:10.1016/J.COGNITION.2021.104758

28. Patenaude B, Smith SM, Kennedy DN, Jenkinson M. A Bayesian model of shape and appearance for subcortical brain segmentation. Neuroimage. 2011;56(3):907–922. doi:10.1016/J.NEUROIMAGE.2011.02.046

29. Smith SM, Zhang Y, Jenkinson M, et al. Accurate, robust, and automated longitudinal and cross-sectional brain change analysis. Neuroimage. 2002;17(1):479–489. doi:10.1006/nimg.2002.1040

30. Morel P. Gramm: grammar of graphics plotting in Matlab. J Open Source Softw. 2018;3(23):568. doi:10.21105/JOSS.00568

31. Lakens D. Calculating and reporting effect sizes to facilitate cumulative science: A practical primer for t-tests and ANOVAs. Front Psychol. 2013;4(NOV). doi:10.3389/FPSYG.2013.00863/ABSTRACT

32. Diedenhofen B, Musch J. cocor: A Comprehensive Solution for the Statistical Comparison of Correlations. PLoS One. 2015;10(4). doi:10.1371/JOURNAL.PONE.0121945

33. DeLong ER, DeLong DM, Clarke-Pearson DL. Comparing the Areas under Two or More Correlated Receiver Operating Characteristic Curves: A Nonparametric Approach. Biometrics. 1988;44(3):837. doi:10.2307/2531595

34. Gorwn B, Carson K. The Basis for Choice Reaction Time Slowing in Alzheimer’s Disease. BRAIN Cogn. 1990;13:148–166.

35. Chen KC, Weng CY, Hsiao S, Tsao WL, Koo M. Cognitive decline and slower reaction time in elderly individuals with mild cognitive impairment. Psychogeriatrics. 2017;17(6):364–370. doi:10.1111/PSYG.12247

36. van Deursen JA, Vuurman EFPM, Smits LL, Verhey FRJ, Riedel WJ. Response speed, contingent negative variation and P300 in Alzheimer’s disease and MCI. Brain Cogn. 2009;69(3):592–599. doi:10.1016/J.BANDC.2008.12.007

37. Andriuta D, Diouf M, Roussel M, Godefroy O. Is Reaction Time Slowing an Early Sign of Alzheimer’s Disease? A Meta-Analysis. Dement Geriatr Cogn Disord. 2019;47(4-6):281–288. doi:10.1159/000500348

