## Supplementary materials for "Performance and validation of a digital memory test across the Alzheimer’s Disease continuum"

| | EHC | SCI | MCI | AD | p-value all | p-value EHC/SCI | p-value SCI/MCI | p-value MCI/AD |
|---|---|---|---|---|---|---|---|---|
| **Age** | 66.6 (7.5) | 59.9 (8.6) | 69.3 (11.5) | 69.8 (11.2) | *0.040 | 0.165 | 0.065 | 1 |
| **Gender (M/F)** | 6/15 | 8/7 | 8/4 | 5/7 | 0.175 | 0.714 | 1 | 0.877 |
| **Education** | 15.8 (3.6) | 15.9 (5.2) | 14.0 (4.7) | 15.5 (6.2) | 0.448 | 1 | 1 | 1 |
| **ACE** | 97.4 (2.0) | 94.3 (3.5) | 86.7 (7.7) | 71.7 (5.2) | *<0.001 | 0.052 | *<0.001 | *<0.001 |
| **DS** | 17.7 (2.9) | 18.2 (3.8) | 17.2 (4.8) | 13.8 (2.2) | *0.007 | 1 | 1 | 0.079 |

**Table S1 | Demographics and tests – longitudinal subset**

| | EHC | SCI | MCI | AD | p-value all | p-value EHC/SCI | p-value SCI/MCI | p-value MCI/AD |
|---|---|---|---|---|---|---|---|---|
| **Age** | 66.6 (7.5) | 57.7 (7.2) | 65.8 (11.3) | 69.2 (9.7) | *<0.001 | *<0.001 | *0.048 | 0.546 |
| **Gender (M/F)** | 24/37 | 14/17 | 4/5 | 22/15 | 0.285 | 0.592 | 0.969 | 0.415 |
| **Education** | 16.2 (3.8) | 16.0 (4.5) | 16.6 (4.4) | 14.1 (3.6) | *0.046 | 1 | 1 | 0.411 |
| **ACE** | 97.2 (2.5) | 93.9 (5.7) | 89.6 (3.9) | 70.5 (19.3) | *<0.001 | 0.335 | 0.335 | *<0.001 |

**Table S2 | Demographics and tests – neuroimaging sample**

P-values are derived by the between-group ANOVA (p-value all) with subsequent Holm post-hoc comparisons (p-value EHC/SCI, p-value SCI/MCI, p-value MCI/AD).

### EFFECTS OF SET SIZE AND DELAY

| | **Set size** | **Delay** | **Set size x Delay** |
|---|---|---|---|
| **Identification Time** | F = 493.6, ***p < 0.001**<br>η2 = 0.466 | F = 122.0, ***p < 0.001**<br>η2 = 0.037 | F = 4.1, ***p = 0.043**<br>η2 = 0.001 |
| **Localization Time** | F = 423.2, ***p < 0.001**<br>η2 = 0.416 | F = 161.8, ***p < 0.001**<br>η2 = 0.046 | F = 1.8, p = 0.186 |
| **Identification Accuracy** | F = 1298.7, ***p < 0.001**<br>η2 = 0.624 | F = 168.2, ***p < 0.001**<br>η2 = 0.043 | F = 20.7, ***p < 0.001**<br>η2 = 0.006 |
| **Absolute Localization Error** | F = 1625.5, ***p < 0.001**<br>η2 = 0.726 | F = 109.5, ***p < 0.001**<br>η2 = 0.019 | F = 35.0, ***p < 0.001**<br>η2 = 0.005 |
| **Target detection** | F = 3088.8, ***p < 0.001**<br>η2 = 0.845 | F = 34.1, ***p < 0.001**<br>η2 = 0.004 | F = 0.008, p = 0.927 |
| **Misbinding** | N/A | F = 0.3, p = 0.577 | N/A |
| **Guessing** | F = 145.0, ***p < 0.001**<br>η2 = 0.211 | F = 96.6, ***p < 0.001**<br>η2 = 0.045 | F = 1.3, p = 0.249 |
| **Imprecision** | F = 49.6, ***p < 0.001**<br>η2 = 0.085 | F = 78.8, ***p < 0.001**<br>η2 = 0.043 | F = 5.0, ***p = 0.001**<br>η2 = 0.002 |

**Table S3 | Cross-sectional analysis of Set size and Delay effects**

Set size (1 vs 3 items), Delay (1 vs 4 seconds), N/A = not applicable.

| | Metric | Set size | Delay | Set size x Delay |
|---|---|---|---|---|
| Group EHC | **Identification Time** | F = 572.5, ***p < 0.001**<br>η2 = 0.641 | F = 118.3, ***p < 0.001**<br>η2 = 0.051 | F = 6.0, ***p = 0.016**<br>η2 = 0.002 |
| | **Localization Time** | F = 580.2, ***p < 0.001**<br>η2 = 0.634 | F = 176.7, ***p < 0.001**<br>η2 = 0.070 | F = 2.4, p = 0.125 |
| | **Identification Accuracy** | F = 664.2, ***p < 0.001**<br>η2 = 0.605 | F = 89.5, ***p < 0.001**<br>η2 = 0.057 | F = 23.9, ***p < 0.001**<br>η2 = 0.013 |
| | **Absolute Localization Error** | F = 827.3, ***p < 0.001**<br>η2 = 0.732 | F = 64.2, ***p < 0.001**<br>η2 = 0.022 | F = 28.5, ***p < 0.001**<br>η2 = 0.009 |
| | **Target detection** | F = 1389.5, ***p < 0.001**<br>η2 = 0.838 | F = 27.6, ***p < 0.001**<br>η2 = 0.006 | F = 3.828, p = 0.052 |
| | **Misbinding** | N/A | F = 6.107, ***p = 0.015**<br>η2 = 0.038 | N/A |
| | **Guessing** | F = 229.0, ***p < 0.001**<br>η2 = 0.444 | F = 50.1, ***p < 0.001**<br>η2 = 0.036 | F = 0.7, p = 0.407 |
| | **Imprecision** | F = 181.6, ***p < 0.001**<br>η2 = 0.370 | F = 49.7, ***p < 0.001**<br>η2 = 0.042 | F = 2.3, p = 0.131 |
| Group SCI | **Identification Time** | F = 125.8, ***p < 0.001**<br>η2 = 0.592 | F = 33.8, ***p < 0.001**<br>η2 = 0.052 | F = 14.7, ***p < 0.014**<br>η2 = 0.014 |
| | **Localization Time** | F = 107.9 ***p < 0.001**<br>η2 = 0.554 | F = 46.6, ***p < 0.001**<br>η2 = 0.071 | F = 14.5, ***p < 0.001**<br>η2 = 0.013 |
| | **Identification Accuracy** | F = 152.5, ***p < 0.001**<br>η2 = 0.590 | F = 20.2, ***p < 0.001**<br>η2 = 0.041 | F = 4.7, ***p = 0.035**<br>η2 = 0.008 |
| | **Absolute Localization Error** | F = 212.8, ***p < 0.001**<br>η2 = 0.732 | F = 9.3, ***p = 0.004**<br>η2 = 0.010 | F = 4.5, ***p = 0.038**<br>η2 = 0.004 |
| | **Target detection** | F = 328.4, ***p < 0.001**<br>η2 = 0.808 | F = 2.2, p = 0.148 | F = 0.000, p = 0.980 |
| | **Misbinding** | N/A | F = 0.1, p = 0.703 | N/A |
| | **Guessing** | F = 58.9, ***p < 0.001**<br>η2 = 0.420 | F = 6.9, ***p = 0.011**<br>η2 = 0.020 | F = 1.1, p = 0.308 |
| | **Imprecision** | F = 38.5, ***p < 0.001**<br>η2 = 0.334 | F = 6.0, ***p = 0.018**<br>η2 = 0.019 | F = 0.6, p = 0.450 |
| Group MCI | **Identification Time** | F = 114.5, ***p < 0.001**<br>η2 = 0.546 | F = 15.0, ***p < 0.001**<br>η2 = 0.019 | F = 1.5, p = 0.222 |
| | **Localization Time** | F = 108.0, ***p < 0.001**<br>η2 = 0.524 | F = 14.3, ***p < 0.001**<br>η2 = 0.022 | F = 0.8, p = 0.385 |

| | | | | |
|---|---|---|---|---|
| | **Identification Accuracy** | F = 409.3, *p < 0.001<br>η2 = 0.730 | F = 27.1, *p < 0.001<br>η2 = 0.030 | F = 1.5, p = 0.218 |
| | **Absolute Localization Error** | F = 589.0, *p < 0.001<br>η2 = 0.730 | F = 10.6, *p = 0.002<br>η2 = 0.009 | F = 4.2, *p = 0.046<br>η2 = 0.003 |
| | **Target detection** | F = 948.7, *p < 0.001<br>η2 = 0.889 | F = 1.0, p = 0.309 | F = 1.784, p = 0.187 |
| | **Misbinding** | N/A | F = 2.8, p = 0.099 | N/A |
| | **Guessing** | F = 27.0, *p < 0.001<br>η2 = 0.201 | F = 20.5, *p < 0.001<br>η2 = 0.070 | F = 0.3, p = 0.560 |
| | **Imprecision** | F = 19.4, *p < 0.001<br>η2 = 0.142 | F = 17.9, *p < 0.001<br>η2 = 0.069 | F = 0.2, p = 0.618 |
| **Group AD** | **Identification Time** | F = 27.1, *p < 0.001<br>η2 = 0.211 | F = 12.0, *p < 0.001<br>η2 = 0.029 | F = 0.7, p = 0.413 |
| | **Localization Time** | F = 21.5, *p < 0.001<br>η2 = 0.164 | F = 16.9, *p < 0.001<br>η2 = 0.033 | F = 0.1, p = 0.748 |
| | **Identification Accuracy** | F =322.2, *p < 0.001<br>η2 = 0.661 | F = 32.8, *p < 0.001<br>η2 = 0.039 | F = 0.4, p = 0.534 |
| | **Absolute Localization Error** | F = 288.6, *p < 0.001<br>η2 = 0.683 | F = 33.4, *p < 0.001<br>η2 = 0.036 | F = 3.0, p = 0.081 |
| | **Target detection** | F = 816.3, *p < 0.001<br>η2 = 0.871 | F = 8.1, *p = 0.006<br>η2 = 0.004 | F = 2.798, p = 0.099 |
| | **Misbinding** | N/A | F = 0.9, p = 0.341 | N/A |
| | **Guessing** | F = 0.6, *p = 0.443<br>η2 = 0.006 | F = 26.6, *p < 0.001<br>η2 = 0.069 | F = 0.9, p = 0.342 |
| | **Imprecision** | F = 2.0, p = 0.001<br>η2 = 0.020 | F = 22.1, *p < 0.001<br>η2 = 0.061 | F = 4.3, *p = 0.042<br>η2 = 0.009 |

**Table S4 | Group-by-group Set size and Delay effects**

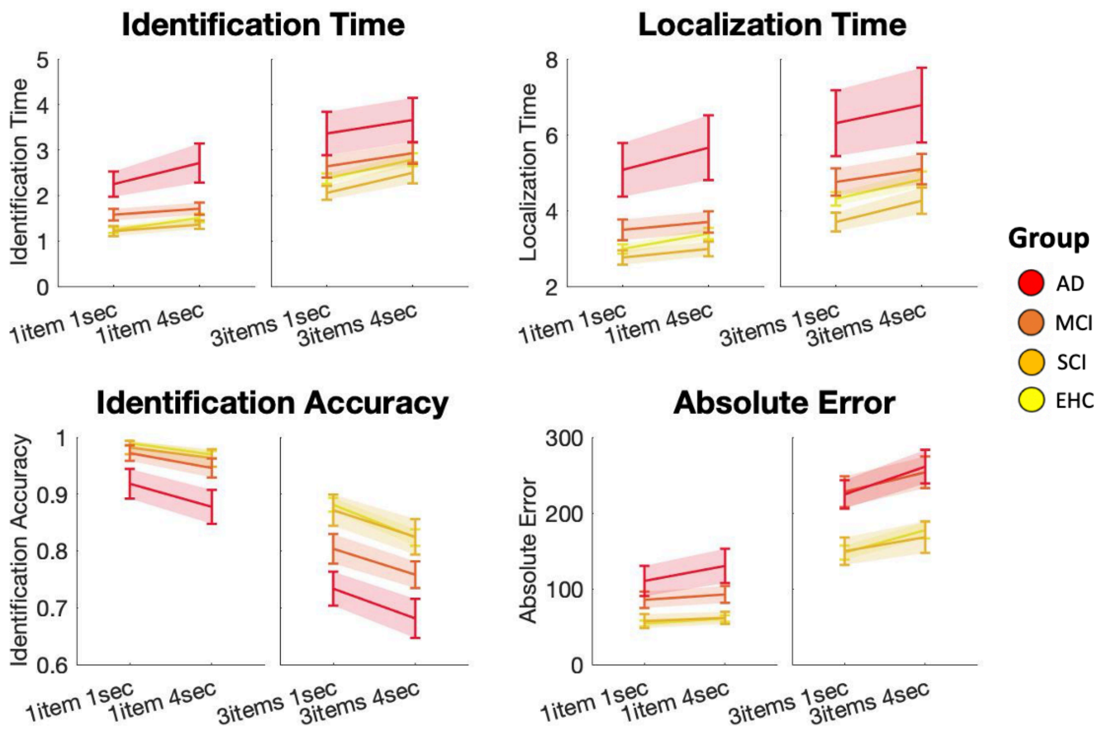


**Figure S1 | Transdiagnostic analysis: Effects of Set Size and Delay-basic metrics**

EHC = yellow, SCI = light orange, MCI = dark orange, AD = red.

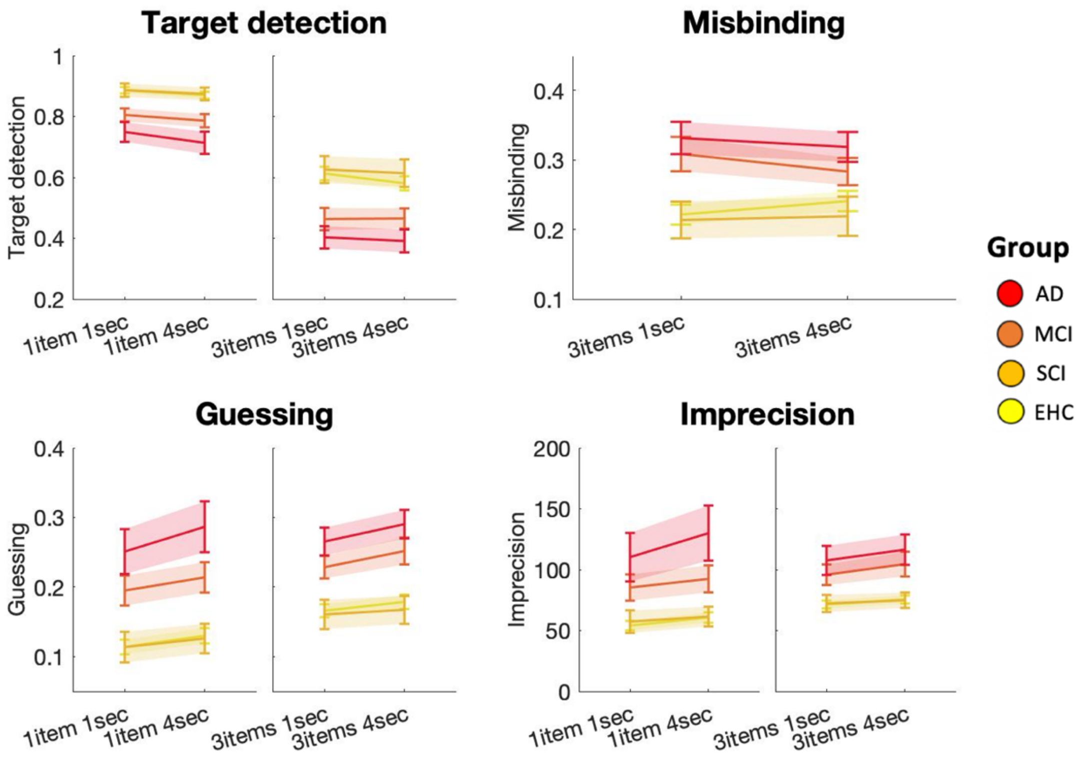


**Figure S2 | Transdiagnostic analysis: Effects of Set Size and Delay- mixture model metrics**

EHC = yellow, SCI = light orange, MCI = dark orange, AD = red.

| | Group | Session | Group x Session |
|---|---|---|---|
| **Identification Accuracy** | F = 46.6, ***p < 0.001**<br>η2 = 0.551 | n.s | n.s |
| **Absolute Localization Error** | F = 64.9, ***p < 0.001**<br>η2 = 0.614 | n.s | F = 3.2, ***p = 0.025**<br>η2 = 0.031 |
| **Identification Time** | F = 35.0, ***p < 0.001**<br>η2 = 0.472 | n.s | n.s |
| **Localization Time** | F = 20.3, ***p < 0.001**<br>η2 = 0.333 | F = 4.3, ***p = 0.039**<br>η2 = 0.024 | n.s |
| **Target detection** | F = 72.7, ***p < 0.001**<br>η2 = 0.667 | n.s | n.s |
| **Guessing** | F = 67.6, ***p < 0.001**<br>η2 = 0.640 | n.s | n.s |
| **Misbinding** | F = 28.9, ***p < 0.001**<br>η2 = 0.447 | n.s | n.s |
| **Imprecision** | F = 45.9, ***p < 0.001**<br>η2 = 0.507 | n.s | F = 4.8, ***p = 0.004**<br>η2 = 0.053 |

**Table S5 | Longitudinal dataset - Group and Session effects**

Session represents baseline session (Time 0) and follow-up session after 1 year. n.s. = not significant.

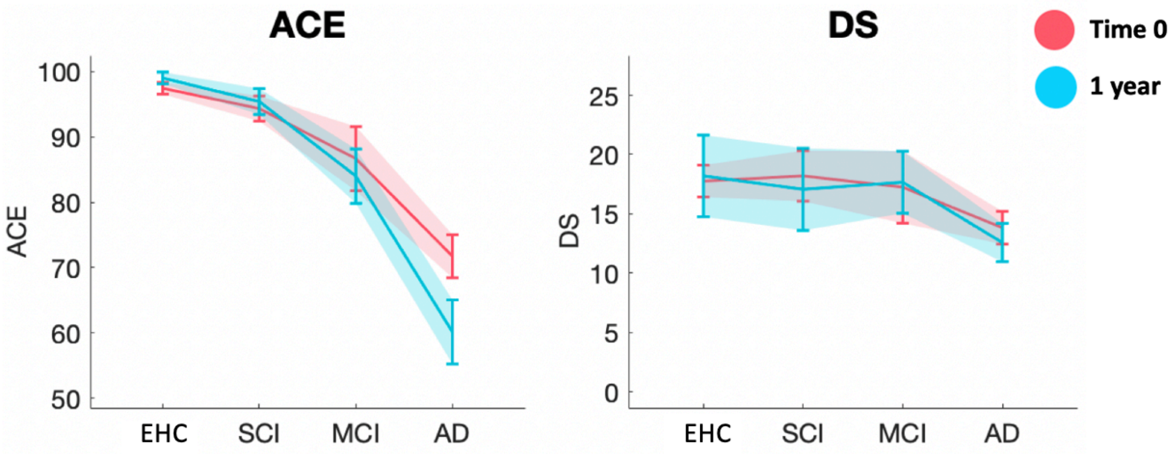


**Figure S3 | Longitudinal analysis of standard neuropsychological tests**

| | **Decline in ACE scores** |
|---|---|
| **Identification time** | F (1,45), t = 4.5, *p = 0.002, $R^2$ = 0.3143 |
| **Localization time** | F (1,45), t = 4.6, *p =0.001, $R^2$ = 0.3239 |
| **Identification accuracy** | F (1,45), t = -5.0, *p < 0.001, $R^2$ = 0.3537 |
| **Absolute Localization Error** | F (1,45), t = 5.3, *p < 0.001, $R^2$ = 0.3537 |
| **Target detection** | F (1,45), t = -5.2, *p < 0.001, $R^2$ = 0.3902 |
| **Misbinding** | F (1,45), t = 4.0, *p < 0.001, $R^2$ = 0.2706 |
| **Guessing** | F (1,45), t = 4.6, *p = 0.005, $R^2$ = 0.3333 |
| **Imprecision** | F (1,45), t = 5.5, *p < 0.001, $R^2$ = 0.4107 |

#### Table S6 | Longitudinal dataset – prediction of cognitive decline
